# A Modelling Study for Designing a Multi-layered Surveillance Approach to Detect the Potential Resurgence of SARS-CoV-2

**DOI:** 10.1101/2020.06.27.20141440

**Authors:** Yang Liu, Wenfeng Gong, Samuel Clifford, Maria E. Sundaram, CMMID-COVID19 Working Group, Mark Jit, Stefan Flasche, Petra Klepac

**Affiliations:** Centre for the Mathematical Modelling of Infectious Diseases, Department of Infectious Disease Epidemiology, London School of Hygiene & Tropical Medicine, London, UK; Bill and Melinda Gates Foundation, Seattle, WA, USA; Rollins School of Public Health, Emory University, Atlanta, GA, USA; School of Public Health, University of Hong Kong, Hong Kong SAR, China; Department for Applied Mathematics and Theoretical Physics, University of Cambridge

**Keywords:** routine surveillance, lab-based surveillance, early warning, non-pharmaceutical interventions, dynamic model, intervention evaluation, testing, COVID-19

## Abstract

**Background:** Countries achieving control of COVID-19 after an initial outbreak will continue to face the risk of SARS-CoV-2 resurgence. This study explores surveillance strategies for COVID-19 containment based on polymerase chain reaction tests.

**Methods:** Using a dynamic SEIR-type model to simulate the initial dynamics of a COVID-19 introduction, we investigate COVID-19 surveillance strategies among healthcare workers, hospital patients, and community members. We estimate surveillance sensitivity as the probability of COVID-19 detection using a hypergeometric sampling process. We identify test allocation strategies that maximise the probability of COVID-19 detection across different testing capacities. We use Beijing, China as a case study.

**Findings:** Surveillance subgroups are more sensitive in detecting COVID-19 transmission when they are defined by more COVID-19 specific symptoms. In this study, fever clinics have the highest surveillance sensitivity, followed by respiratory departments. With a daily testing rate of 0.07/1000 residents, via exclusively testing at fever clinic and respiratory departments, there would have been 598 [95% eCI: 35, 2154] and 1373 [95% eCI: 47, 5230] cases in the population by the time of first case detection, respectively. Outbreak detection can occur earlier by including non-syndromic subgroups, such as younger adults in the community, as more testing capacity becomes available.

**Interpretation:** A multi-layer approach that considers both the surveillance sensitivity and administrative constraints can help identify the optimal allocation of testing resources and thus inform COVID-19 surveillance strategies.

**Funding:** Bill & Melinda Gates Foundation, National Institute of Health Research (UK), National Institute of Health (US), the Royal Society, and Wellcome Trust.

## Introduction

Coronavirus disease 2019 (COVID-19) is an infectious disease caused by the severe acute respiratory syndrome coronavirus 2 (SARS-CoV-2) and was first detected in Wuhan, China towards the end of 2019.^1^ On 11 Mar 2020, the World Health Organisation declared COVID-19 a global pandemic.^2^ Within six months of its emergence, COVID-19 has led to over eight million reported cases and over 450,000 reported deaths globally.^3^ Many countries and regions have succeeded in reducing COVID-19 incidence after the initial epidemics.

Nevertheless, it is unlikely that SARS-CoV-2 will be eradicated in the near future. Given the high transmissibility of the pathogen,^4,5^ the non-trivial proportion of infectious individuals showing mild to no symptoms,^6^ the non-specific nature of symptoms,^7^ the highly intertwined global travel network, and a lack of effective pharmaceutical measures for prevention or therapy,^8^ countries that successfully contain the initial spread of COVID-19 will likely continue to face risks introduced by international travellers and unidentified local cases. With physical distancing measures gradually easing, sporadic infection clusters have already been observed.^9^

To prevent these sporadic infection clusters from seeding new epidemics, rapid infection detection is vital. Containment strategies, such as case isolation and contact tracing, can quickly be overwhelmed if transmission remains undetected for too long. Thus, sustainable, cost-effective, and highly sensitive surveillance systems for SARS-CoV-2 are essential to the success of COVID-19 containment.

This study explores different surveillance systems that maximise the probability of COVID-19 detection using polymerase chain reaction (PCR) while minimising the material and human resources required. We consider a hypothetical city with the population size, age structure, and healthcare infrastructure similar to that of Beijing, China. Fever clinics, a triage system that emerged during the 2003 outbreak of SARS-CoV, play a crucial role in COVID-19 response in China^10^ and could be considered a potentially effective surveillance option elsewhere. The framework introduced is relevant to containment strategies in countries exiting the initial phases of COVID-19 epidemics, where only a small number of cases are observed sporadically.

## Methods

### The Epidemic Process

We simulate the spread of SARS-CoV-2 using a deterministic age-stratified compartmental SEIR-type model (Figure 1).^11^ Additionally, the infectious compartment is split into *I*^*pc*^, *I*^*c*^ and *I*^*sc*^ to account for differences in disease progression. The compartment *I*^*c*^ represents infectious individuals whose symptoms are sufficiently severe (“clinical” illnesses) for them to seek healthcare; *I*^*pc*^ represents the pre-clinical infectious individuals who have not yet developed symptoms; *I*^*sc*^ represents infectious individuals who may not seek healthcare for COVID-19 due to mild symptoms (“subclinical” illnesses). This framework has been used to study COVID-19 in several previous studies.^6,12,13^ More details about this model can be found in the Supplemental Material Section 1.

**Figure 1.**
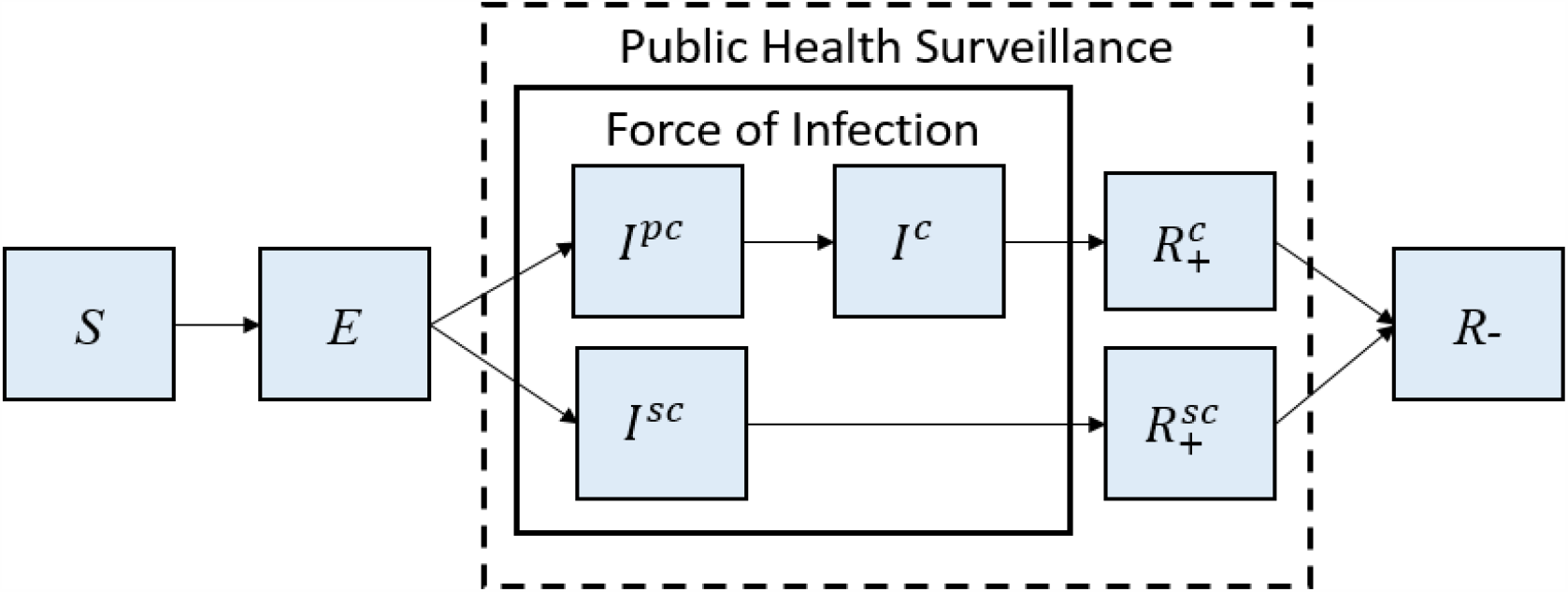
Conceptual Diagram of the COVID-19 Dynamic Model. *S*: Susceptible; *E*: Exposed; *I*^*pc*^: Pre-clinical Infectious; *I*^*c*^: Clinical Infectious; *I*^*sc*^: Sub-clinical Infectious; 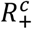: PCR-positive Removed following *I*^*c*^; and 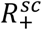: PCR-positive Removed following *I*^*sc*^; *R*_−_: PCR-negative Removed. Solid box indicates compartments that affect the force of infection; dashed box indicates compartments detectable by PCR-based surveillance.

Chau et al. discovered that COVID-19 patients may remain PCR positive for up to 10 days after hospital admission, at which point they may no longer be infectious. ^14^ To accounts for this, building upon the existing model structure, we break the R compartment (“Removed”, a state in which individuals are no longer able to transmit illness) down into 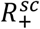 (PCR-positive individuals from *I*^*sc*^),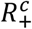 (PCR-positive individuals from *I*^*c*^) and *R*_−_(PCR-negative individuals). This study assumed perfect specificity.^15^ Potential false-positive results may be compensated by double-testing positive samples.

We obtained other model parameters from the literature or online databases (Table 1). At the start of the simulation, we assume there was one exposed younger adult (i.e., 15-64 year-old); the population is otherwise susceptible, consistent with low seropositivity found in serological surveys.^16,17^ A meta-analysis in Davies et al. showed an unmitigated reproduction number of COVID-19 to be 2.7 (95% critical interval: 1.6-3.9).^12^ Here, we assume an effective reproduction number (R_t_) of 2 that reflects a 25% reduction from 2.7 as a result of public health measures (e.g., physical distancing in public). Additionally, R_t_ of 2.7 (unmitigated) and 1.4 (50% reduction) are explored as possible scenarios. At each R_t_ level, sensitivity analyses given different sets of epidemiological parameters are provided, with results presented as Uncertainty Range (UR).

**Table 1.** Parameters Table.

| Parameters | Baseline Scenario | Sensitivity Analyses | Source |
| --- | --- | --- | --- |
| **Epidemiological parameters** |  |  |  |
| Latent period | 2.6 days | 1 - 4 days | Li et al.<sup>1</sup>, Backer et al.<sup>18</sup>, and Davies et al.<sup>12</sup> |
| Pre-clinical infectious period | 2.4 days | 1 - 3 days | Liu et al.<sup>19</sup> and He et al.<sup>20</sup> |
| Clinical infectious period | 3.6 days | 3 - 6 days | He et al.<sup>20</sup> |
| Subclinical infectious period (assumed to be the same as the sum of pre-clinical and clinical infectious periods) | 6 days | 5 and 8 days | Davies et al.<sup>12</sup> |
| Age-specific clinical fraction (relative that | 0.28 - 0.69 | - | Davies et al.<sup>6</sup> |
| develop symptoms after infection) |  |  |  |
| Relative susceptibility | 0.44 - 1<br>(Baseline group: 60-69 year-olds) | - | Davies et al.<sup>6</sup> |
| Relative transmissibility of subclinical infections (compared to clinical) | 0.14 | 0.5 | Keeling et al.<sup>21</sup> and Davies et al.<sup>12</sup> |
| Mitigated reproduction number | 2 | 1.4, 2.7 | Assumed based on Davies et al.<sup>12</sup> |
| Number of contacts between individuals of different ages | 3 - 20 contacts/ day | - | Prem et al.<sup>13</sup> |
| Population (counts and age distribution) | 549,734 - 2,686,112 / age group | - | China Statistics Yearbook<sup>22</sup> |
| **Surveillance parameters** |  |  |  |
| Prevalence of influenza-like illness (ILI) | 3% | 2%, 6% | Chinese National Influenza Weekly Reports<sup>23</sup> |
| Annual prevalence of all respiratory illnesses<sup>^</sup> | ~10% | - | GBD 2017 Disease and Injury Incidence and Prevalence Collaborators<sup>24</sup> |
| Maximum routine testing capacity per population-day | 4.15/1000 |  | Wan<sup>25</sup> |
| Daily Fever Clinic Service Capacity, per population-day | 0.07/1000 |  | Gong<sup>26</sup> |
| Daily Respiratory Department Service Capacity, per population-day | 1.35/1000 | 0.81-1.89/1000 | Zhang et al.<sup>25</sup> |
| Healthcare worker to patient ratio | 1:3 | - | Chinese National Influenza Weekly Report,<sup>23</sup> Zhang et al.,<sup>27</sup> China Health Statistics Yearbook<sup>28</sup> |
| PCR sensitivity for detection of SARS-CoV-2 | Clinical: 30%~90%<br>Subclinical: 18%~46% | - | Chau et al.<sup>14</sup> |
| PCR specificity for detection of SARS-CoV-2 | 100% | - | Grassly et al.<sup>15</sup> |

<sup>^</sup> Respiratory illnesses are defined as “respiratory infections and tuberculosis” (excluding latent tuberculosis infections), “chronic respiratory diseases”, and “Tracheal, bronchus, and lung cancer”.

### Surveillance Layers and Subgroups

Surveillance layers are settings where COVID-19 surveillance is possible; surveillance subgroups are defined by specific characteristics (e.g., age and occupation) within a given surveillance layer (Table 2). Fever clinics are triage systems established in China during the 2003 SARS epidemic to limit the spread of pandemic pathogens in hospital settings and allow rapid detection.^29^ Telephone-based triaging systems combined with drive-through testing centres seen during the COVID-19 pandemic are motivated similarly. In the context of COVID-19, anyone with any respiratory symptoms and potential exposure to a confirmed case, as well as anyone with a combination of fever and any respiratory symptoms but no known exposure, are encouraged to present to fever clinics in China.^30^

**Table 2.** Surveillance Pathways.

| Surveillance Layers | Surveillance Subgroups | Descriptions | HCW |  |  |  |  | Non-HCW |  |  |  |  |
| --- | --- | --- | --- | --- | --- | --- | --- | --- | --- | --- | --- | --- |
| | | | $I^{pc}$ | $I^c$ | $R_+^c$ | $I^{sc}$ | $R_+^{sc}$ | $I^{pc}$ | $I^c$ | $R_+^c$ | $I^{sc}$ | $R_+^{sc}$ |
| Healthcare Worker Surveillance | Fever Clinic | HCW working in different hospital departments | x |  |  | x | x |  |  |  |  |  |
|  | Respiratory Department |  | x |  |  | x | x |  |  |  |  |  |
|  | Other Hospital Departments |  | x |  |  | x | x |  |  |  |  |  |
| Patient-based Surveillance | Fever Clinic | Patients seeking care with influenza-like symptoms |  | x |  |  |  |  | x |  |  |  |
|  | Respiratory Department | Patients seeking care for any respiratory conditions |  | x |  |  |  |  | x |  |  |  |
|  | Other Hospital Departments | Patients seeking care for any non-respiratory conditions | x |  | x | x | x | x |  | x | x | x |
| Community-based Surveillance | Children | $\leq 15$ years | x | x | x | x | x | x | x | x | x | x |
|  | Younger Adults | 16 - 64 years | x | x | x | x | x | x | x | x | x | x |
| | Older Adults | $\geq 65$ years | x | x | x | x | x | x | x | x | x | x |

This study further assumes 1) that individuals with clinical respiratory illness who seek care would be captured by a surveillance system in either fever clinics or respiratory departments; 2) that those with a subclinical COVID-19 infection who are seeking care for non-respiratory causes may be detected in other hospital departments; 3) that those with a COVID-19 infection who do not actively seek care may be detected in the community whether they exhibit symptoms or not. Specific pathways to the detection of different surveillance layers and subgroups are shown in Table 2.

The risk of a susceptible individual, whether an HCW or not, contracting COVID-19 while present in a fever clinic or respiratory department is elevated due to the concentration of infectious individuals. HCWs may face a higher COVID-19 infection risk than patients due to prolonged exposures. In this study, the expected number of infectious encounters for HCW and non-HCW adults are calculated by taking the product of the number of contacts (either patient or non-patient, digitised from Jiang et al.^31^) and the corresponding COVID-19 prevalence (inpatient or non-patient settings). The HCW to non-HCW ratios of infectious encounters (*α*) is then used as a time-varying risk multiplier for different HCW subgroups. More details can be found in the Supplemental Material Section 2.

### Surveillance Sensitivity and Probability of Detection

The surveillance sensitivity (*φ*_*i,k,t*_) is the probability of detecting at least one individual infectious with COVID-19 in a subgroup *i* on a given day *t* (since the first case) is characterised by a hypergeometric distribution:

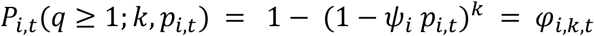

where *k* is the number of individuals tested on day *t, p*_*i,t*_ is the probability of an individual in subgroup *i* on day *t* testing positive, *q* is the number of infectious individuals identified through testing and *Ψ*_*i*_ is the sensitivity of PCR in subgroup *i*. In this study, *p*_*i,t*_ depends on the surveillance pathways outlined in Table 2. For example, for a patient seeking care at a fever clinic, the probability that they are infectious with SARS-CoV-2 is:

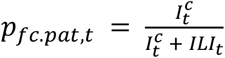

where *ILI*_*t*_ is the number of non-COVID-19 patients who could be triaged into fever clinics based on influenza-like illness (ILI). This equation does not include *I*^*pc*^ and *I*^*sc*^ as these compartments would not be triaged into a fever clinic by definition.

The probability of an individual in subgroup *i* on day *t* testing positive (*p*_*i,t*_) decreases as the background population size increases (under the assumption of perfect specificity). In the example above, the background population in a fever clinic is captured by *ILI*_*t*_. In respiratory departments, the background population is the number of patients who visit respiratory departments for all non-COVID-19 respiratory causes (*Rsp*_*t*_). Thus, at any given time *t, Rsp*_*t*_ must be at least as large as *ILI*_*t*_. The proportion of COVID-19-infected patients in fever clinics *p*_*fc*.*pat,t*_ is always larger than that in respiratory departments (*p*_*rsp*.*pat,t*_) by definition.

We assume that HCWs who have clinical respiratory illness behave identically to other non-HCW individuals with clinical respiratory illness, and hence will attend fever clinics or respiratory departments. Surveillance targeting the HCW subgroups will only capture those current in the *I*^*pc*^, *I*^*sc*^, or 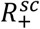 (see Table 2). These HCWs will face a higher risk of infection due to occupational exposure (*α*_*fc*.*hcw,t*_). For example, the probability of an HCW working at a fever clinic to be PCR detectable on a given day *t* can be expressed as:

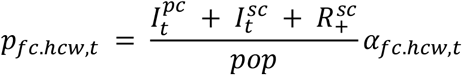

where *α*_*fc*.*hcw,t*_ is the increased risk multiplier for an HCW working at a fever clinic on day *t*. The probability *π*_*t*_ of detecting at least one COVID-19 infection by time *t* and its associated modelled infection prevalence (clinical, subclinical or both) based on at least one subgroup can be expressed as:

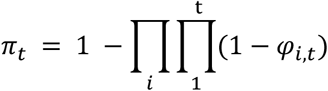

### Comparing Fever Clinics and Respiratory Departments

The testing capacity needed at the respiratory departments to achieve a similar probability of COVID-19 detection while testing at maximum capacity at fever clinics (1,600 tests/day or 0.07 tests per thousand residents per day (t/k/day)) is found using mean absolute errors. Then at different levels of daily testing capacity, we estimate the time it takes for the outbreak to be detected by simulating 10,000 surveillance processes, using *π*_*i,t*_ as the probability of a binomial process. The cumulative incidences (i.e., those who are no longer in the *S* compartment) are then extracted to approximate the scope of the underlying epidemic size when the outbreak was detected. The relevant uncertainty is expressed using the middle 95% of the simulated sample, i.e., the 95% empirical confidence intervals (eCI).

### Characteristics of Surveillance Systems

In practice, surveillance systems are constrained by human and capital resources as well as individuals available in the target subgroup (driven by local healthcare infrastructure and demographics (Table 1). In terms of testing capacity, Wuhan could test up to 6 t/k/day by 22 April 2020.^32^ South Korea and the United Kingdom are testing at 1 to 3 t/k/day levels.^33,34^ We cap the maximum daily testing capacity at 4.15t/k/day (or 100,000 tests for Beijing), consistent with the observed testing capacity in Beijing.^25^ Additionally, we assume a maximum of 10% HCWs at non-fever clinic hospital departments, and 30% of HCWs at fever clinics can be tested each day as a part of HCW surveillance to minimise interference with every-day work.

### Determining the most efficient strategy

We consider a wide range of testing capacities between 0 and 100,000 tests daily, with increments of 200 tests, totalling 501 different levels of testing capacity. Therefore, the number of daily tests available at level *n* is 200*(n-1)*. We then determined how these 200*(n-1)* tests should be allocated across different subgroups to maximise the probability of detection (*π*_*t*_). Instead of an exhaustive search among all possible allocations of resources, we used a recursive algorithm. Assuming there are only two subgroups, *i* and *j*, the probability of detecting COVID-19 transmission can be expressed as:

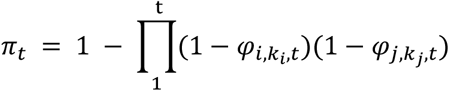

where the sum of *k*_*i*_ and *k*_*j*_ is *(n* − *1)*. Then, in the next step, 200*n* tests can be best allocated as:

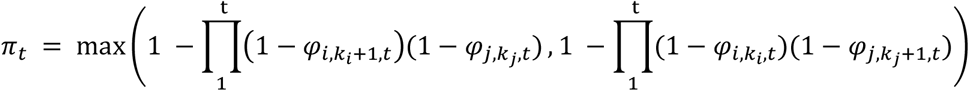

where the sum of *k*_*i*_and *k*_*j*_ + 1and of *k*_*i*_ + 1and *k*_*j*_are both *n*. This recursive algorithm is applied to all nine surveillance subgroups to identify the optimal surveillance strategies, i.e., the allocation of COVID-19 tests that can achieve the highest *π*_*t*_.

All analyses were done in R. Code used is publicly available [https://github.com/yangclaraliu/covid_surveillance_strategy].

## Results

### The subgroups-specific surveillance sensitivity

Surveillance sensitivity is the probability that on-going COVID-19 transmission (i.e., one or more cases) can be detected on a given day *t*. Based on the assumption of an emerging yet undetected epidemic, the surveillance sensitivities of different subgroups are shown in Figure 2. At the baseline scenario (R_t_ = 2), it took 20 days for the epidemic process to incur over 100 cumulative incidences. It took 17 and 25 days using unmitigated or 50% reduced R_t_ to reach this cumulative incidence level. At this point in the outbreak, while conducting 1,600 tests per day, the probability that a surveillance strategy exclusively targeting fever clinic patients and HCW can detect more than one COVID-19 case is 1.1% [UR: 0.6%, 1.3%] and 1.2%[UR: 1.0%, 2.2%], respectively. At the same daily testing level, the probability of detection is only 0.5%[UR: 0.3%, 0.6%] and 0.7%[0.4%, 0.9%], respectively, for respiratory departments patients and HCW. Among age-specific community groups, conducting surveillance among younger adults yields the highest surveillance sensitivity (0.4% [UR: 0.3%, 0.4%]). When other respiratory pathogens are common and the number of respiratory patients per day is high, patient-based surveillance performs relatively worse than its annual average (Supplemental Material Section 3).

**Figure 2.**
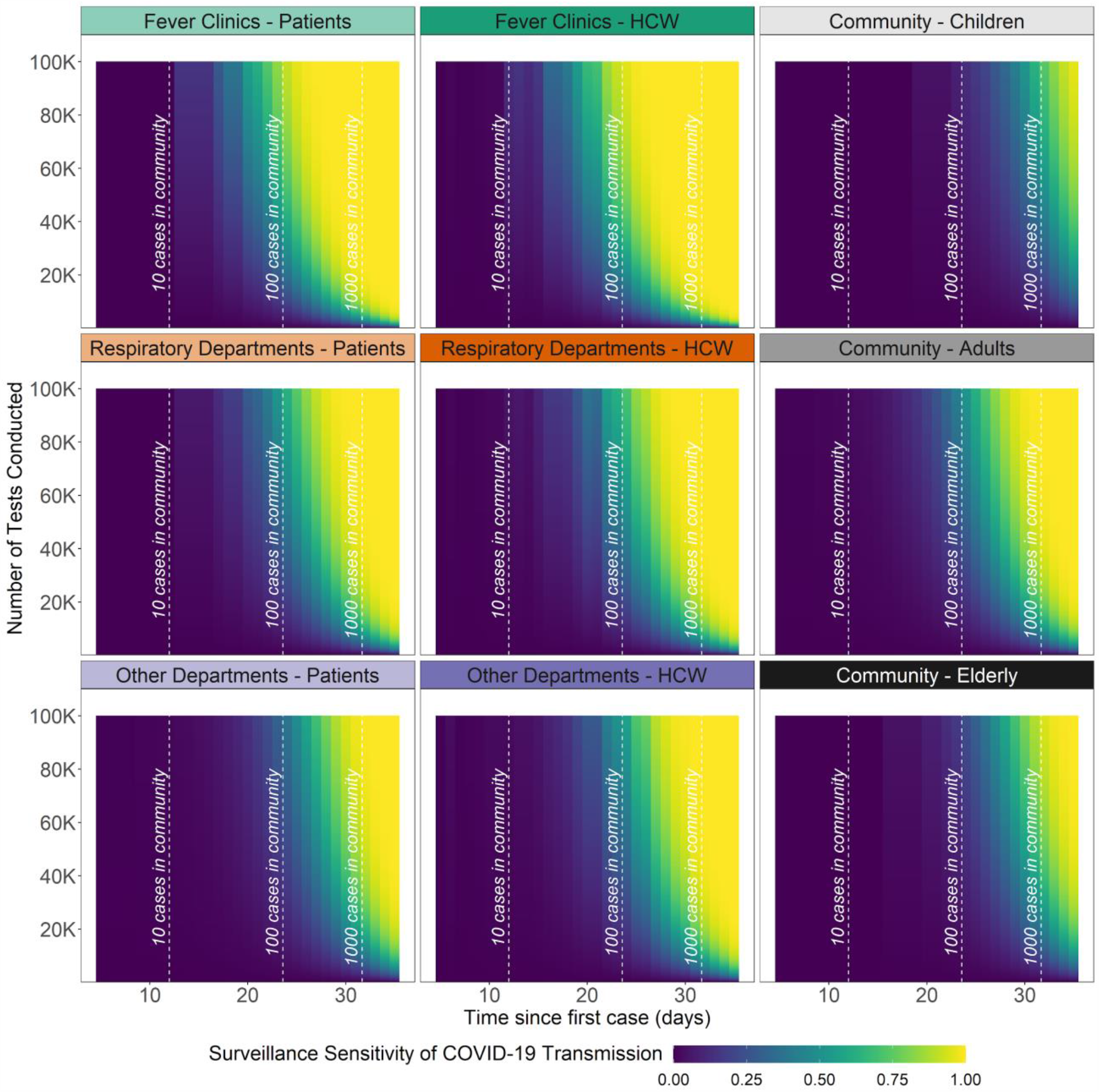
**Surveillance sensitivity (*φ*_*i,k,t*_)** is the probability of detecting at least one COVID-19 case on day *t* after the first infected case (i.e., seeding event) in a surveillance subgroup had a certain amount of daily testing been conducted. Vertical dashed lines represent thresholds of >10, >100, and >1000 cases in the population. Healthcare-related administrative constraints (e.g., number of HCWs available) are not yet incorporated into this output.

### Comparing Fever Clinics and Respiratory Departments

To further understand the potential benefits of fever clinics, we compared the probability of detecting at least one case of COVID-19 by day *t* using realistic administrative constraints (e.g., fever clinic service capacity) for the city of Beijing, China. Using the parameters outlined in Table 1, we estimated that the maximum service capacity at fever clinic is 1,600 patients/day, respiratory departments 29,400 patients/day, and other hospital departments more than a quarter-million patients/day. If there is a capacity to conduct 1,600 tests/day (i.e., 0.07 t/k/day), the surveillance system was able to reach a 50% chance of detection among fever clinic patients 3 days earlier than among respiratory department patients. Using an unmitigated *R*_*t*_ shrunk the time gained to 2 days; a 50% reduced *R*_*t*_ of 1.4 expanded time gained to 4 days. To achieve an equivalent COVID-19 detection performance, patient-based surveillance in respiratory departments would need more than double the rate of testing (i.e. 3,600 tests/day, 0.15 t/k/day).

When testing all patients arriving at respiratory departments (i.e., 29,400 tests per day, 1.22 t/k/day), we estimate a 12.3% [UR: 4.2%, 20.4%] and 25.2% [UR: 9.3%, 37.4%] probability to detect at least one COVID-19 case before there were a total of 50 and 100 cases in the simulated epidemic (Figure 3). While testing at maximum capacity in fever clinics (i.e., 1,600 tests per day), there is only a 1.6% [UR: 0.5%, 2.8%] and 3.0% [UR: 1.0%, 5.7%] probability of detecting at least one COVID-19 case before cumulative incidence reaches 50 and 100 cases.

**Figure 3.**
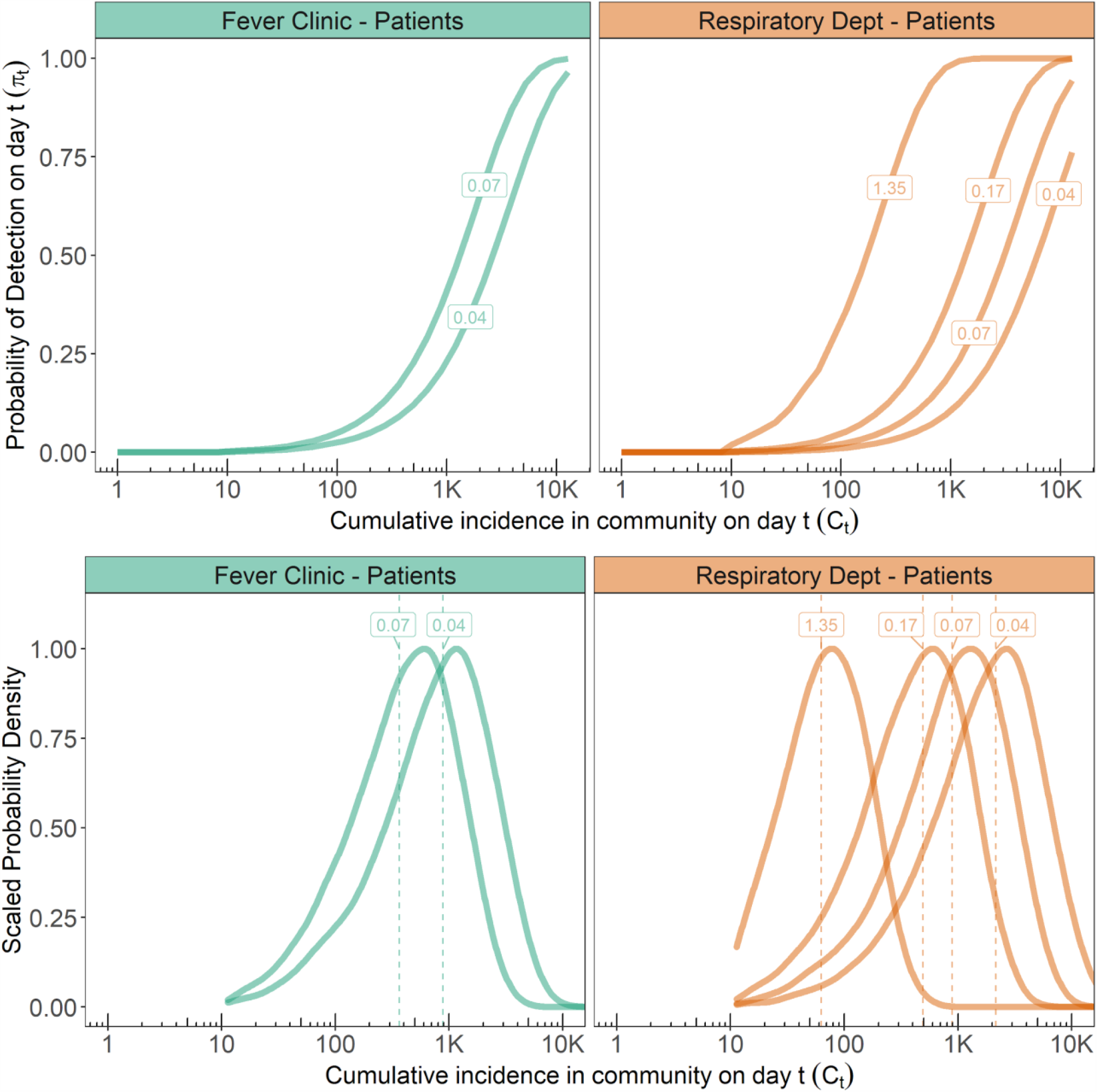
Top: **Probability of detection (***π*_***t***_) at different daily testing capacities with healthcare infrastructure and operational constraints at fever clinics and respiratory departments by day *t*. Bottom: **Empirical probability density functions of cumulative incidence by the time of first COVID-19 case detection** among different surveillance subgroups and given different daily testing capacities. Labels show different daily rates (tests per thousand residents per day, t/k/day).

Using the baseline scenario, we found that while testing at maximum capacity at fever clinics (0.07 t/k/day), the cumulative incidence has likely reached nearly 598 cases [95% eCI: 35, 2154] by the time first COVID-19 is detected. With the same amount of test, the cumulative incidence has reached 1373 cases [95% eCI: 47, 5230] when the first COVID-19 is picked up in fever clinics. Testing at maximum capacity at respiratory departments (1.22 t/k/day) means by the time the first case is detected, the scope of the underlying outbreak is only 91 [95% eCI: 15, 273]. More information about these distributions is included in the Supplemental Material section 4.

### Optimal Strategies at Different Testing Capacity

Figure 4 shows the highest probability of COVID-19 detection achievable at different daily testing capacities. These are also referred to as efficiency frontiers as no higher probability of COVID-19 detection can be achieved without improving the testing capacity. The probability of COVID-19 detection increases as cumulative incidence increases and as daily testing capacities increase. As expected, when cumulative incidence is already high, a small number of tests will already allow us to detect on-going transmission. When daily testing capacities increase, we observe a diminishing return on investment. The additional increase in the probability of detecting COVID-19 by every additional 200 tests conducted decreases as overall daily testing capacity increases. Distinct turning points on the efficiency frontiers indicate when the surveillance system switches from conducting more tests in a given subgroup to testing a new subgroup due to exhausting available individuals.

**Figure 4.**
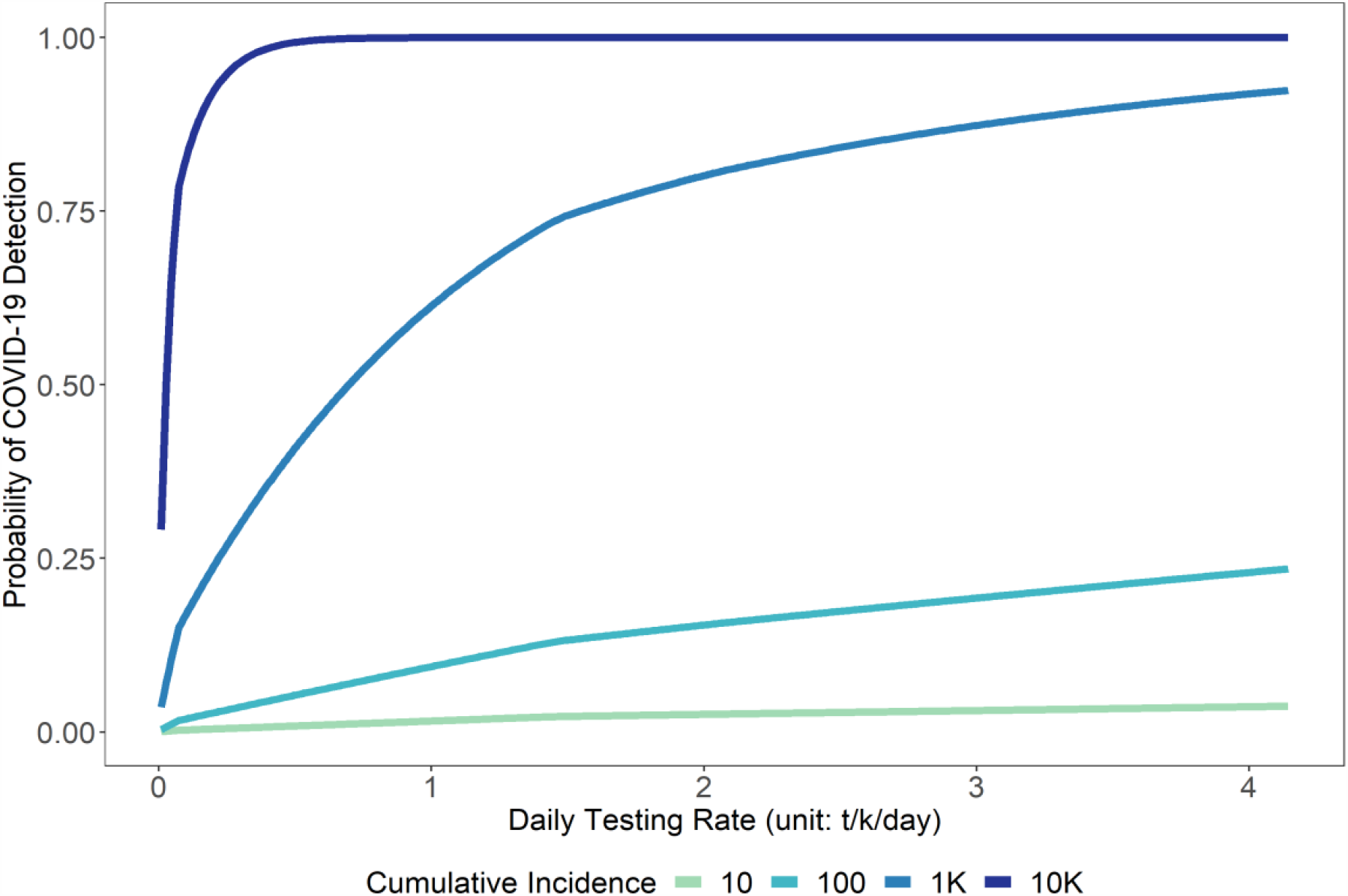
Efficiency frontiers show the optimal strategies at different levels of daily testing capacity and cumulative incidence. The optimal strategy is defined by the maximum probability of COVID-19 detections while considering all possible surveillance layers and subgroups at a given daily testing capacity. The transmission of COVID-19 is considered detected if one or more surveillance layers and subgroups detect at least one infectious case.

Figure 5 shows the specific composition of optimal surveillance strategies along the efficiency frontiers in Figure 4. The surveillance of COVID-19 with the purpose of early detection should prioritise testing among fever-clinic patients and HCWs. When more resources become available, respiratory department patients and HCWs should be included in the surveillance strategy. When more than 30,000 daily testing capacity is reached in a city with 24 million residents, testing among HCWs in other hospital departments and younger adults in the community should be included in the surveillance strategy. During conditions where cumulative incidences are extremely low (i.e., <=100), including these two groups earlier on can improve the probability of detection (see also Supplemental Material Section 5). Resources allocated should be allocated in a stratified manner due to the potential clustering of healthcare-seeking behaviour (See Supplemental Material Section 6).

**Figure 5.**
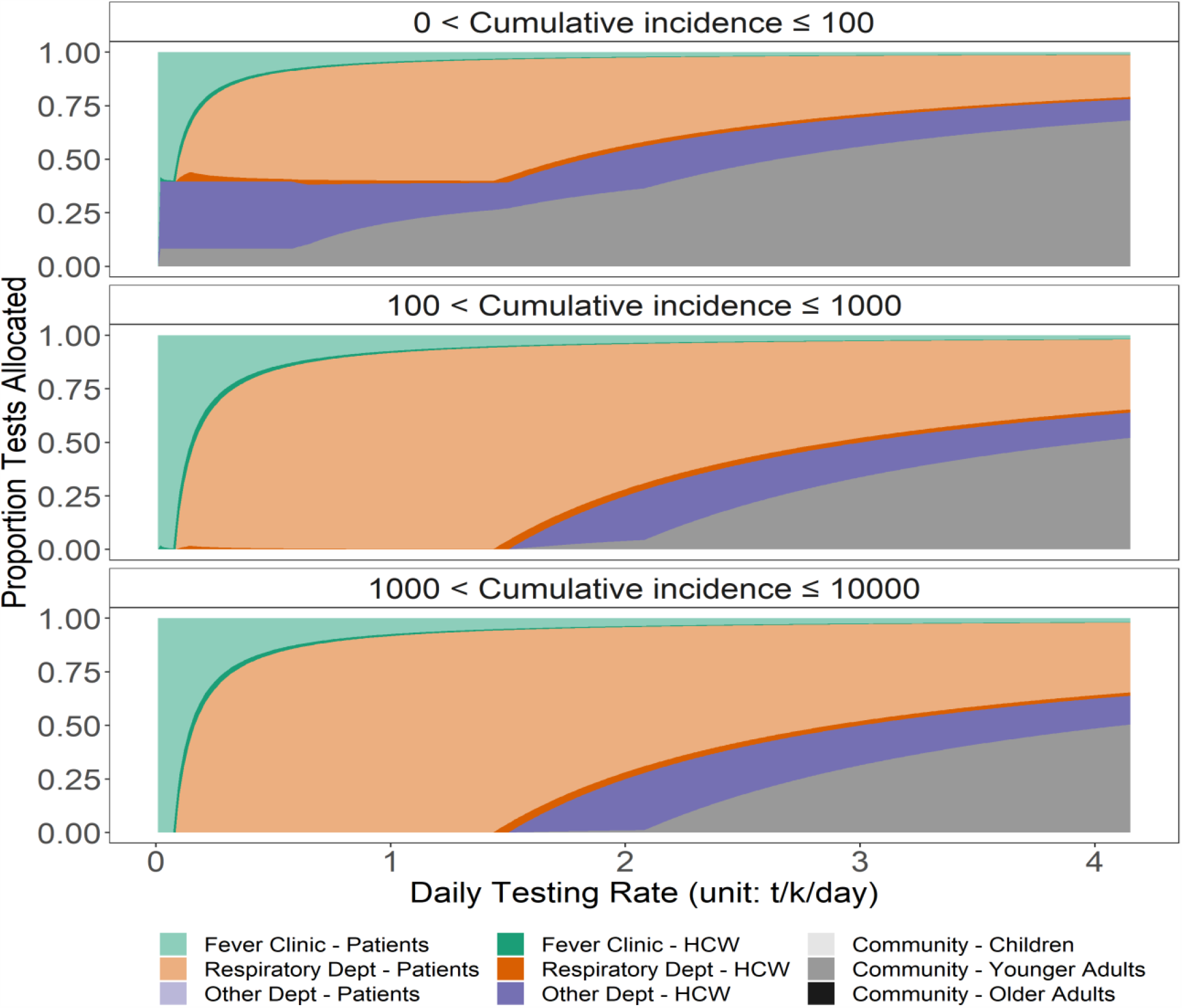
Allocation of resources (by proportion) to different surveillance subgroups along the efficiency frontiers.

## Discussion

This study evaluated different allocations of resources for designing COVID-19 surveillance strategies using Beijing as a case study. We discovered surveillance sensitivity is highest at fever clinics, followed by respiratory departments, and then by community-based subgroups. Testing at maximum capacity among fever clinics patients (i.e., 1,600 tests/day, 0.07 t/k/day) allows us 50% probability to detect on-going transmission by day 29 after the first infection given *R*_*t*_ of 2 (25 when *R*_*t*_ of 2.7, 35 when *R*_*t*_ of 1.4). To achieve similar levels of efficiency, a surveillance system based in the respiratory department would need more than twice the daily testing capacity. Testing at maximum capacity in respiratory patients (29,400 tests/day, 1.35 t/k/day) allows us a 50% probability to detect on-going transmission a week earlier. Surveillance sensitivity is slightly higher among HCWs compared to patients in hospitals given a within-year average condition. However, when other respiratory pathogens are less common, surveillance among fever clinic and respiratory department patients become significantly more sensitive.

The surveillance system in this study is particularly relevant to locations where epidemic “suppression” strategies (e.g., school and workplace closures combined with testing and contact tracing) have been successful. Although countries and regions may experience extended periods without any new COVID-19 cases detected, epidemic risks will continue due to the non-trivial proportion of subclinical cases,^6^ importation from other locations, and the potentially short-lived immunity following infection.^35^ As more countries head into the post epidemic peak phases, surveillance and early detection will continue to play a central role in COVID-19 response. The framework in this study is easily transferable to studying other health systems.

The concepts of fever clinics and respiratory departments should be interpreted broadly as triaging systems, each with a different baseline population defined by different sets of symptoms of various degree of syndromic specificity, leading to different surveillance sensitivity to detect COVID-19 cases. An internal medicine department, for example, is less sensitive compared to the respiratory department but more sensitive than an oncology department.

We are aware of additional triaging criteria based on the assessment of dyspnea, hypoxia, and chest x-ray interpretation.^10^ These criteria may further decrease the background populations for fever clinics and respiratory departments, increasing surveillance sensitivity and thus reducing the number of tests needed. In Mao et al.,^36^ based on only expert assessment and (when needed) chest x-ray, fever clinics in Shanghai were able to triage non-COVID-19 cases with 100% accuracy. This level of triaging accuracy, however, is not always possible. A COVID-19 transmission cluster in Harbin, China, included a patient who was triaged to be non-COVID-19, admitted for cerebral stroke, and thus not isolated. This patient, directly and indirectly, infected 35 persons including three HCWs while admitted.^37^

Routine testing in the community (other hospital departments or age-specific community groups) is less likely to yield a high probability of detecting infections but can be added if early detection is prioritised, in which case we find that the highest probability for detection is among younger adults. Although many studies have found older adults to be the most susceptible age group with the highest clinical fraction who therefore are most likely to seek healthcare, younger adults are likely at a similar risk of infection but may have a smaller probability of displaying symptoms and hence may otherwise go undetected.^1,6,38^

An important assumption in this study is that clinically infectious COVID-19 patients would seek medical care at the same rate as patients with other respiratory pathogens. For example, if 10% of patients infected with other respiratory pathogens seek care, then 10% of COVID-19 patients will seek care. However, this assumption may not be valid. Clinically, COVID-19 patients may be more likely to seek care due to pandemic awareness. They may also be less likely to seek care due to uncertainty around associated healthcare costs or the risk perception surrounding healthcare settings. Public messaging that motivates people to use healthcare may increase the sensitivity of hospital-based COVID-19 surveillance among fever clinics and respiratory departments patients.

As population-level prevalence decreases, the positive predictive value of any individual test will necessarily be lowered, assuming that despite the high specificity of the PCR there will be a small amount of false-positive tests. For example, in an environment completely free of SARS-CoV-2, conducting 30,000 tests/day may still lead to 3 false-positive test results if the specificity is 99.99%. Hence, as testing capacity increases, almost but not quite a perfect testing specificity may lead to false alarms. This will need to be accounted for when deciding how to act upon identified COVID-19 cases. In practice, imperfect test specificity (resulting from factors such as laboratory contamination), could be compensated for by second-testing of the patient or sample.

Highly sensitive COVID-19 surveillance contributes to timely outbreak detection, thereby enabling a swift and targeted response associated with a high probability of containment. In this study, we assessed the surveillance sensitivity in different surveillance layers and subgroups. While designing a COVID-19 surveillance system, prioritisation of fever clinics over respiratory departments and of patients over HCWs tend to optimise for higher COVID-19 detection probability given limited testing capacity.

Community-based testing that targets non-respiratory patients and HCWs or age-specific community groups can capture subclinical infections but may only be considered when more testing capacity becomes available. Future research may focus on assessing the value of information on COVID-19 surveillance systems by estimating the on-ward transmission prevented.

## Data Availability

This study is based on data in the published literature or publicly available online databases.

## Contributors

YL, WFG, PK conceived the idea and designed the study. YL, PK, SC, and SF designed the model with MES and MJ providing input. All coauthors interpreted the results, contributed to writing, and approved the final version for submission.

## Declaration of interests

The authors declare no competing interests.

## Supplemental Material

### 1. The Epidemic Model

The specific structure of the deterministic age-stratified compartmental SE3I3R model in the main text considers five-year age categories for 0-74 year-olds and a single group for everyone above 75 years of age. The specific mechanism shown in Figure 1 is described by the following equations:

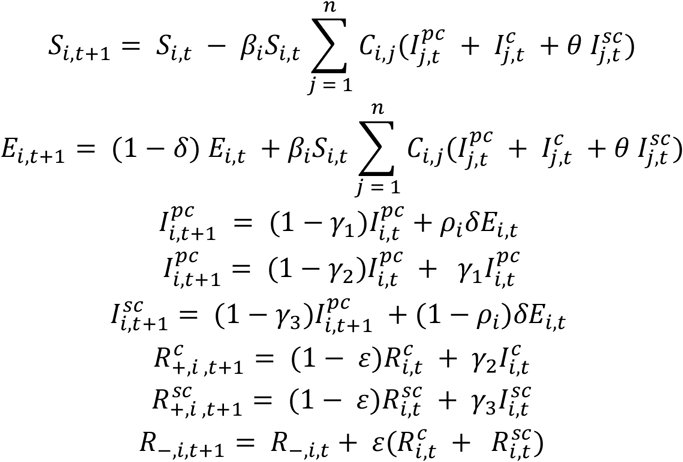

**Table S1.**
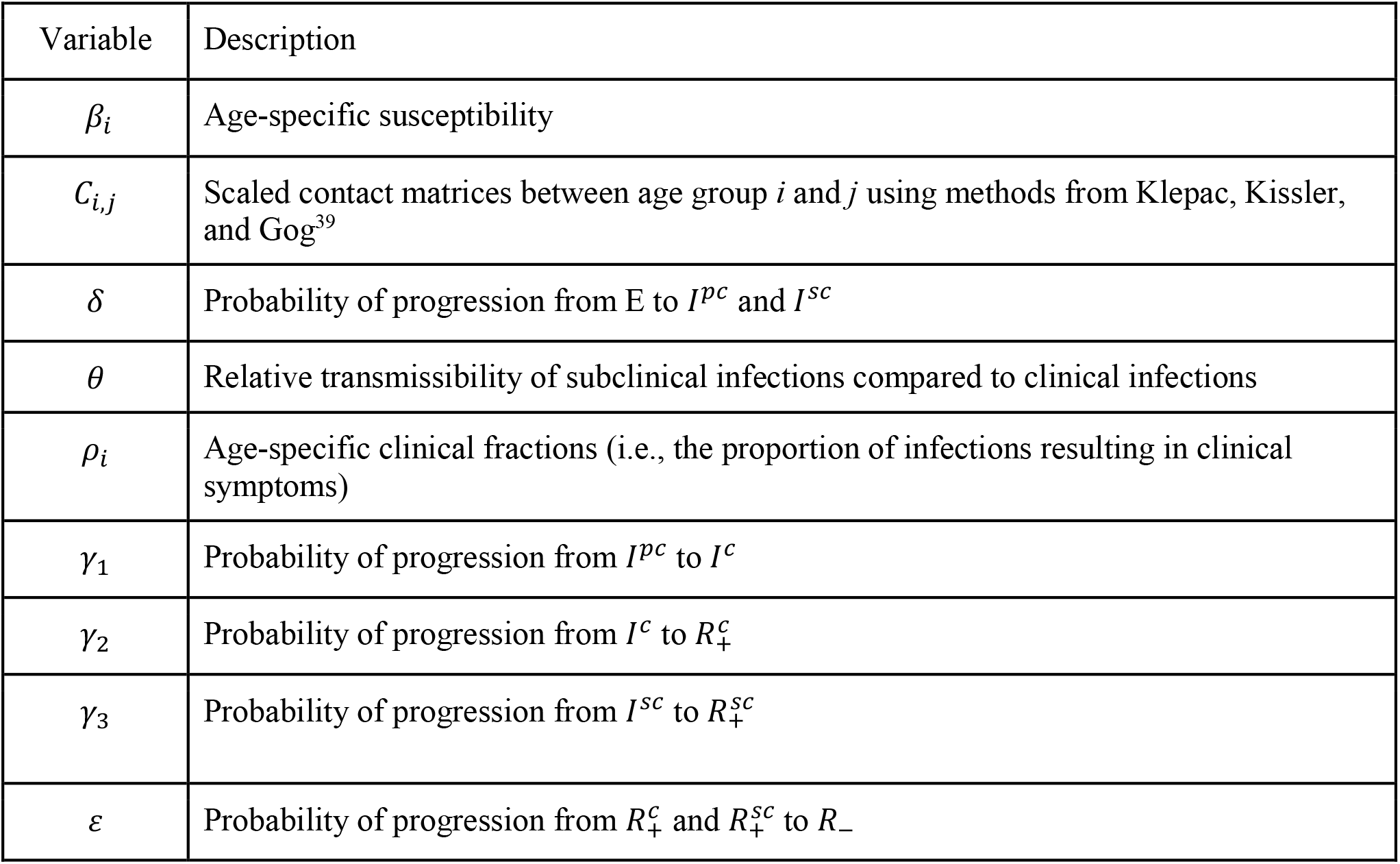
Reference table for symbols used in the equations.

| Variable | Description |
| --- | --- |
| $\beta_i$ | Age-specific susceptibility |
| $C_{i,j}$ | Scaled contact matrices between age group $i$ and $j$ using methods from Klepac, Kissler, and Gog<sup>39</sup> |
| $\delta$ | Probability of progression from E to $I^{pc}$ and $I^{sc}$ |
| $\theta$ | Relative transmissibility of subclinical infections compared to clinical infections |
| $\rho_i$ | Age-specific clinical fractions (i.e., the proportion of infections resulting in clinical symptoms) |
| $\gamma_1$ | Probability of progression from $I^{pc}$ to $I^c$ |
| $\gamma_2$ | Probability of progression from $I^c$ to $R_+^c$ |
| $\gamma_3$ | Probability of progression from $I^{sc}$ to $R_+^{sc}$ |
| $\varepsilon$ | Probability of progression from $R_+^c$ and $R_+^{sc}$ to $R_-$ |

Probability of event (*p*) is calculated using the duration of the event (*d*):

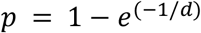

**Figure S1.**
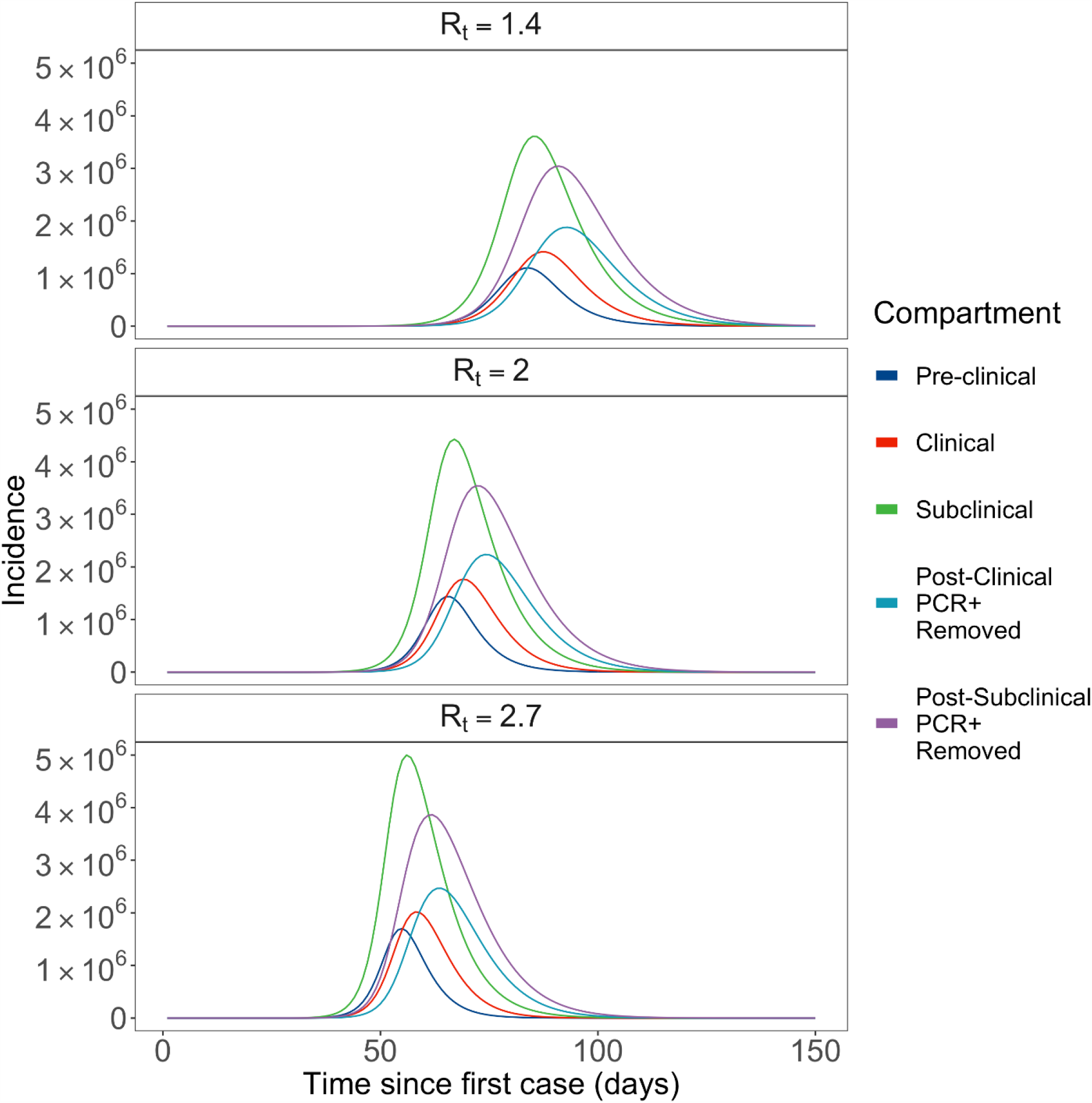
Epidemic curves by PCR detectable surveillance compartments given different reproduction numbers.

### 2. Infection Risk Differential Between Healthcare Workers and General Adults

Marginal probabilities of daily contacts among healthcare workers and adults by setting (i.e., workplace, household, and other) were digitized from Jiang et al.^31^ The study has found a significantly greater number of workplace-related contacts among healthcare workers compared to other adults. In this study, we assumed that the excess contacts are with patients while the remainder is with colleagues. After fitting Poisson distributions and obtaining the respective, we found the expected value of the ratio of workplace contacts between adults and healthcare workers using the following approximation from:^40^

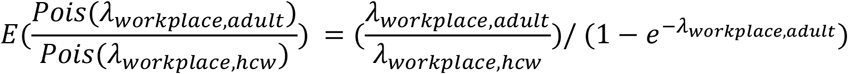

The results show that approximately 35% of workplace contacts by healthcare workers are with colleagues, and 65% are with patients. Using this relationship, we simulated a 1,000 samples for healthcare workers and younger adults, respectively from their empirical distributions shown in Jiang et al.^47^ For a non-HCW younger adult *i* (*i* ∈ {1,2,3…1000}) at time *t*:

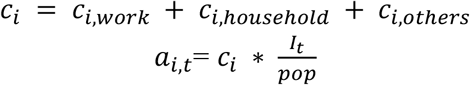

where *c*_*i*_ is the simulated number of contacts, *I* is the total number of infectious individuals (*I*^*pc*^, *I*^*c*^, and *I*^*cc*^) within a population of size *pop*. For a non-HCW younger adult *i* (*i ∈* {1, 2, 3… 1000}) at time *t*:

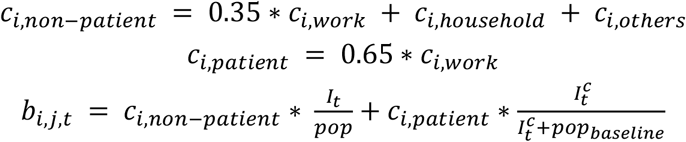

Where 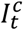 is the number of clinically infectious individuals at time *t* and *pop*_*baseline*_ is the baseline population at a healthcare setting *j*. Using the used in the main text, the increased risks among HCW working in fever clinics can be expressed as:

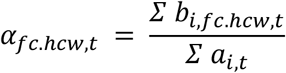

The mean ratio of expected infectious encounters for all three groups of HCW and non-HCW adults are then shown in Figure S2. Assuming equal probabilities for contacts to lead to infection, the ratio of expected infectious encounters can be used to approximate relative infection risks.

**Figure S2.**
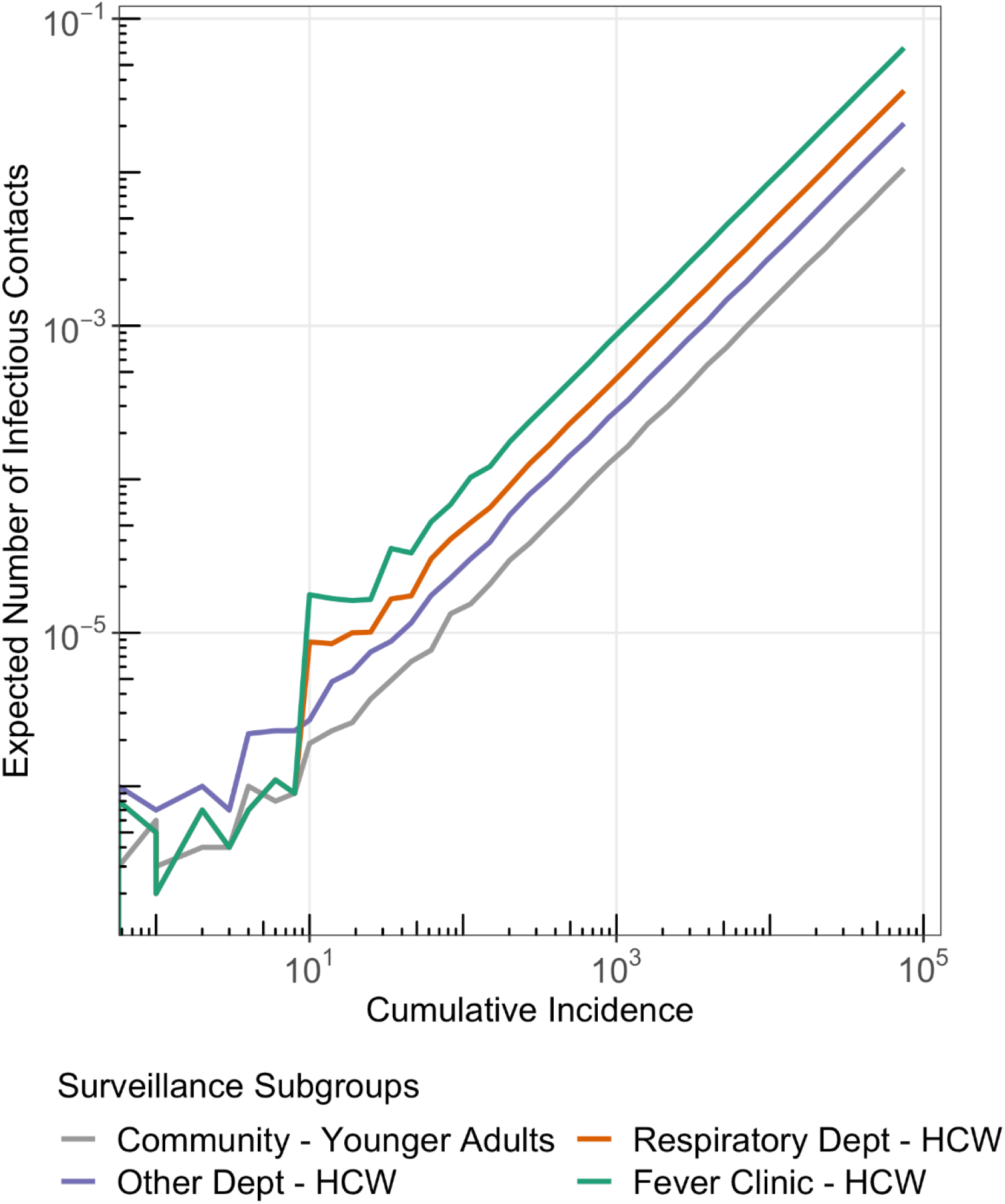
Expected number of encounters with a COVID-19 infectious individual among HCWs in fever clinics, respiratory departments, and other hospital departments, and younger adults in the community.

**Figure S3.**
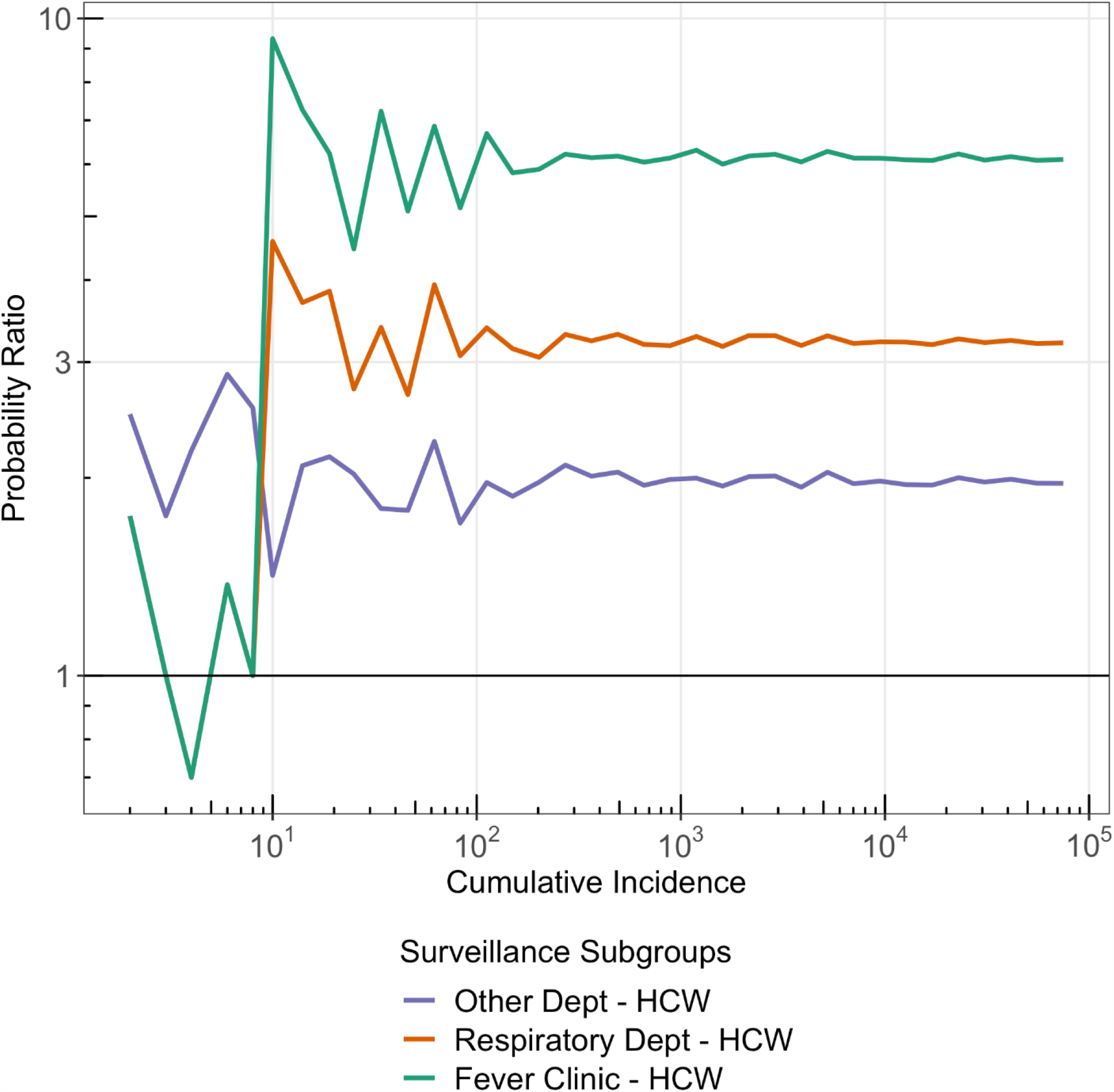
Relative encounters with a COVID-19 infectious individual of HCWs working in different hospital departments compared to younger adults in the community.

### 3. Sensitivity Analysis: Background Healthcare Seeking Rates at Fever Clinics and Respiratory Departments on Probability of COVID-19 Detection

Given the R_t_ of 2 and the baseline set of epidemiological parameters outlined in Table 1 of the main text.

**Table S1.** Scenario a: Baseline scenario, the proportion of ILI among all outpatients and respiratory department service utilisation are both at its annual average. Scenario b: Other respiratory pathogens that often lead to ILI have become less common due to COVID-19 related non-pharmaceutical interventions. Scenario c: During northern hemisphere summer, non-SARS-CoV-2 respiratory pathogens become less common and respiratory department utilisation reaches its annual low. Scenario d: During northern hemisphere winter, non-SARS-CoV-2 respiratory pathogens become more common and respiratory department utilisation reaches its annual high.

| The proportion of ILI among all outpatients | Respiratory Department Service Utilisation | Scenario | Probability of COVID-19 Detection in Patient-based Surveillance |  |  |
| --- | --- | --- | --- | --- | --- |
|  |  |  | Fever Clinic | Respiratory Department | Other Departments |
| 0.03 | 1.35/1000 | a | 1.1 | 0.3 | 0.2 |
| 0.02 | 1.35/1000 | b | 1.6 | 0.4 | 0.2 |
| 0.02 | 0.81/1000 | c | 1.6 | 0.4 | 0.2 |
| 0.06 | 1.89/1000 | d | 0.5 | 0.2 | 0.2 |

### 4. Scope of Epidemics when First Case Has Been Detected

**Table S2.** Characteristics of empirical probability distribution functions of expected epidemic sizes by the time the first case is detected while exclusive testing among fever clinic or respiratory department patients at different levels.

| Daily Testing Capacity |  | Surveillance Subgroup | 2.5 %ile | 25 %ile | Mean | Median | 75 %ile | 97.5 %ile |
| --- | --- | --- | --- | --- | --- | --- | --- | --- |
| Number | Rate (t/k/day) |  |  |  |  |  |  |  |
| 800 | 0.04 | Fever Clinic - Patients | 84 | 888 | 2172 | 2154 | 3891 | 9447 |
| 1,600 | 0.07 | Fever Clinic - Patients | 35 | 203 | 598 | 366 | 888 | 2154 |
| 800 | 0.04 | Respiratory Dept - Patients | 84 | 888 | 2172 | 2154 | 3891 | 9447 |
| 1,600 | 0.07 | Respiratory Dept - Patients | 47 | 366 | 1373 | 888 | 2154 | 5230 |
| 3,600 | 0.17 | Respiratory Dept - Patients | 35 | 203 | 621 | 492 | 888 | 2154 |
| 29,400 | 1.35 | Respiratory Dept - Patients | 15 | 35 | 91 | 63 | 113 | 273 |

### 5. Distribution of Resources Across Surveillance Subgroups

**Figure S4.**
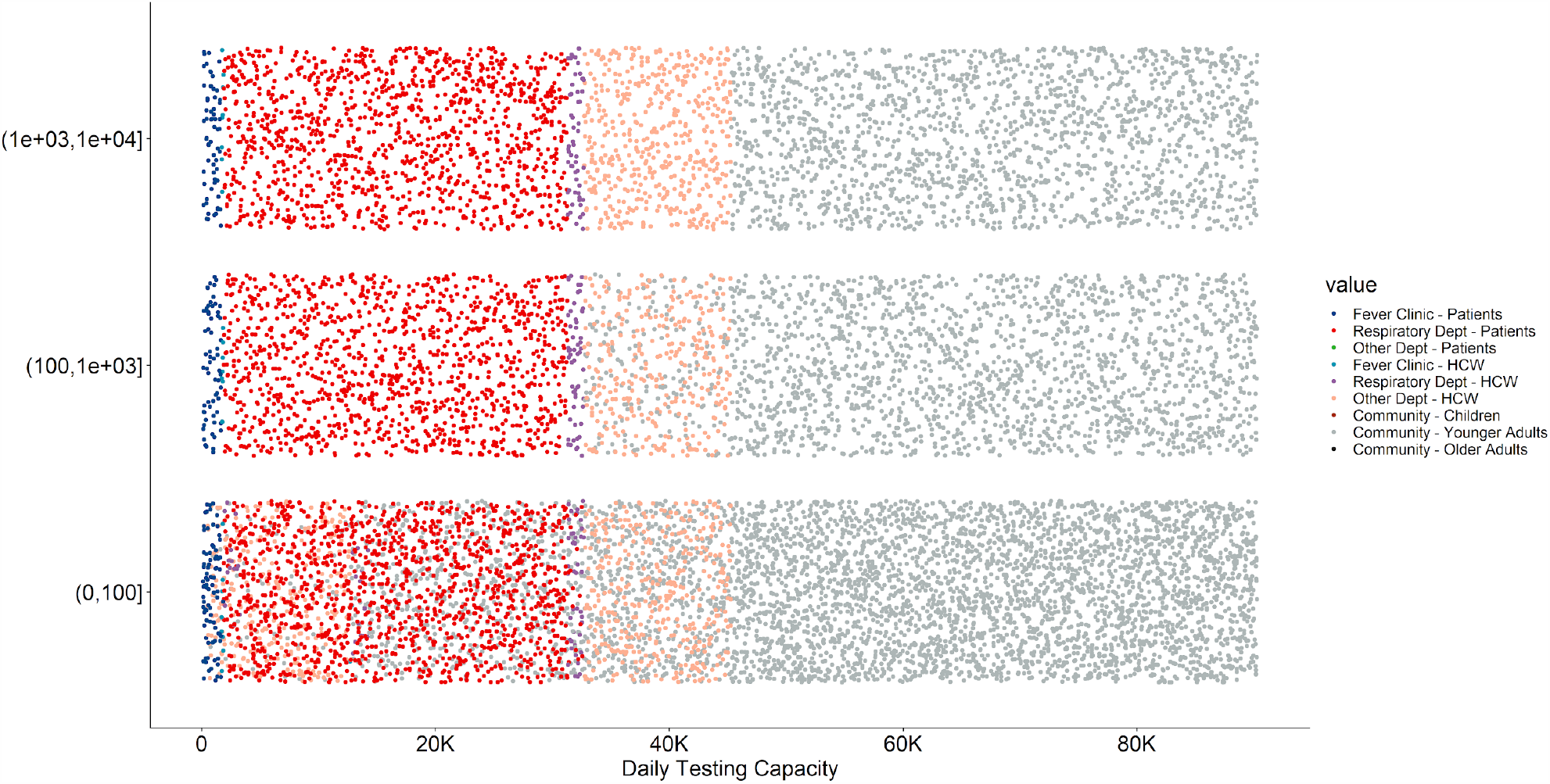
A different presentation of the data shown in Figure 5 of the main text. Y-axis shows three different levels of epidemic sizes. Within each level, the relative position along the y-axis is random and used for visualisation purpose only. At different daily testing capacity levels (shown by the x-axis), the colours represent the decisions made by the recursive algorithms regarding the allocation of the next 200 tests. When cumulative incidence is still low (given y-axis = (0, 100]), including other hospital departments HCWs and younger adults from the general community may be beneficial as they are chosen by the recursive algorithms at lower daily testing capacities.

### 6. The Clustering Effect in Healthcare Seeking Behaviors

The surveillance processes in this study currently assume clinically infectious COVID-19 individuals are equally likely to use all healthcare service options available to them. In reality, at the beginning of an outbreak, healthcare-seeking behaviours are likely clustered. Thus, the potentially detectable HCWs (preclinical, subclinical, and post-subclinical) may also be clustered in a few healthcare providers instead of eventually spread out the entire healthcare system across the city.

We used fever clinic HCWs as an example to explore the potential impacts of clustered healthcare-seeking behaviours on HCW surveillance. The city of Beijing currently runs 79 fever clinics (*n*_*fc*_).^49^ In the main text, we have already described the probability of a given fever clinic HCW to be PCR detectable at time *t* can be expressed as:

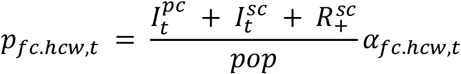

An implicit step that was not covered is:

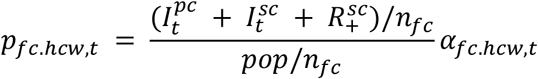

where detectable and the baseline populations are both divided into their catchment unit. To investigate the impact of clustering, we modify the equation above to:

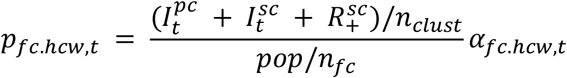

where *n*_*clust*_ is the number of fever clinics that received infectious COVID-19 patients. Results based on *n*_*cluster*_ ∈ {1, 2,…10} with 9%-45% HCWs tested daily are shown below.

We discovered that the optimal number of tests identified in this study should be allocated in a stratified manner. Compared to randomly testing 30% of all fever clinic HCWs, testing a randomly 30% of HCWs in each fever clinic is slightly more efficient, especially when cumulative incidence is low.

**Figure S5.**
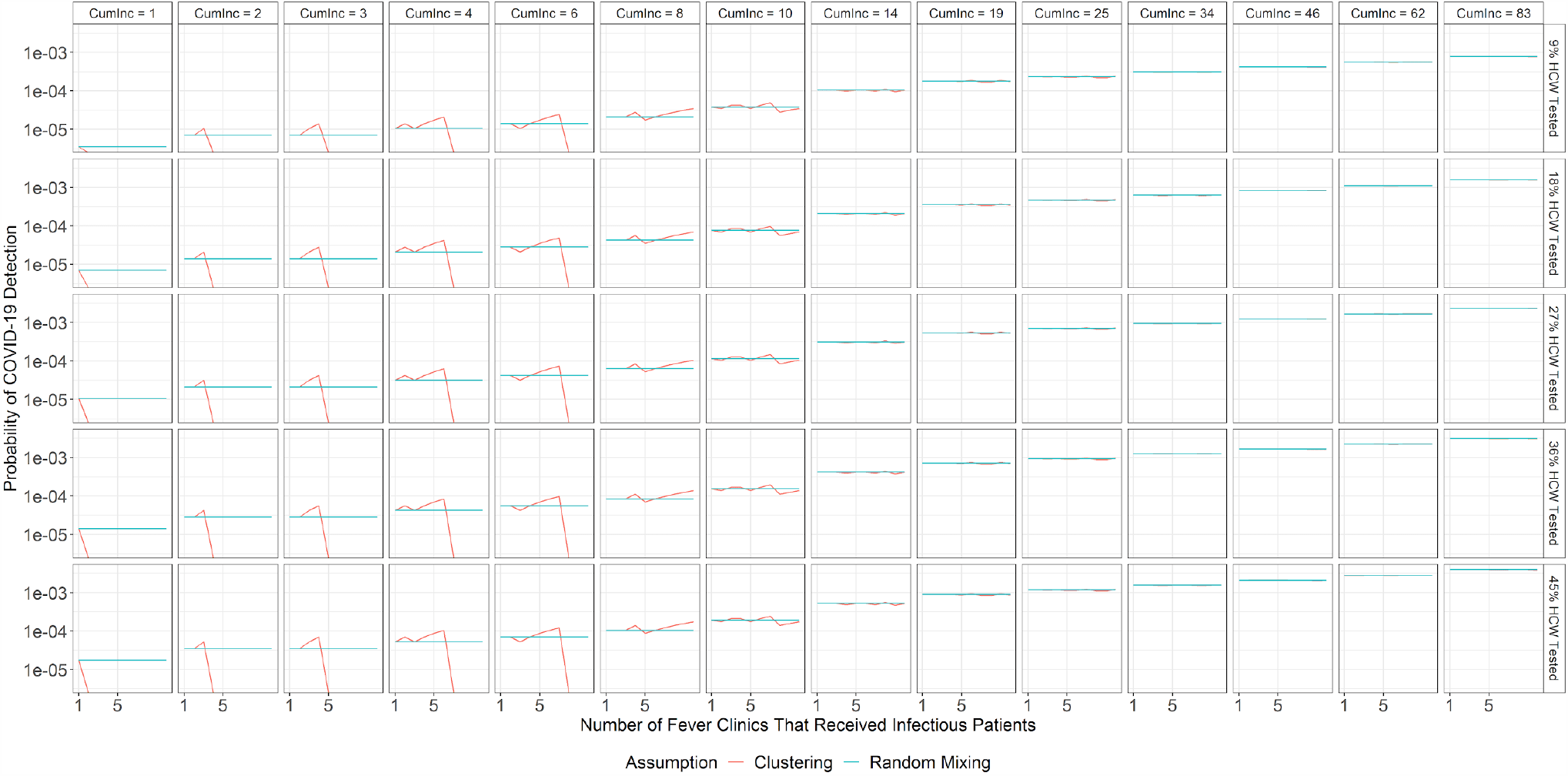
Probability of detecting COVID-19 among HCWs in fever clinics while assuming clustered vs. evenly distributed healthcare seeking behaviours.

**Figure S6.**
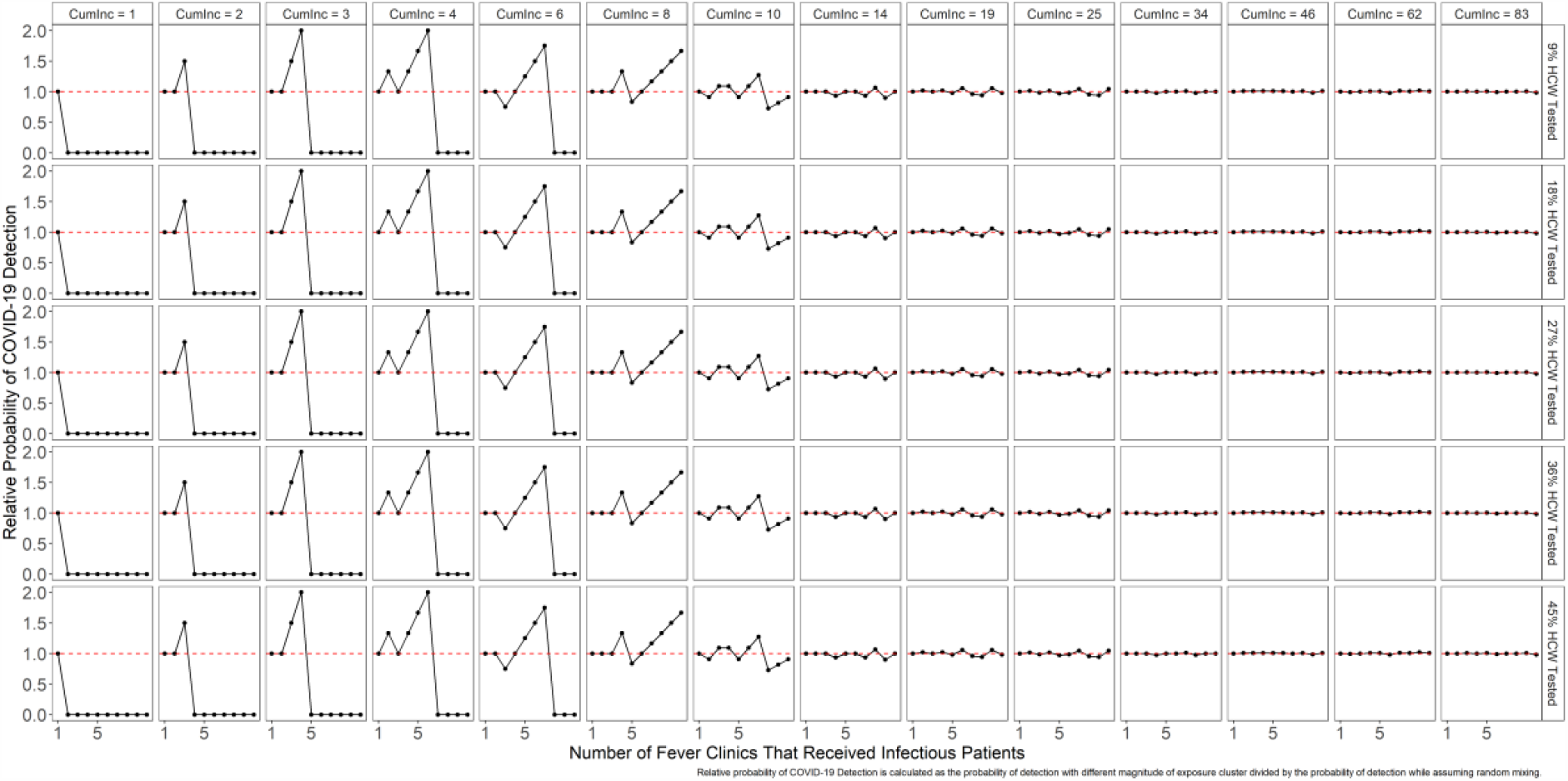
The ratio of the probabilities of detecting COVID-19 among HCWs in fever clinics while assuming clustered over evenly distributed healthcare seeking behaviours. When the magnitude of clustering is high (*n*_*clust*_is small) and the cumulative incidence is low, stratified surveillance is more efficient than surveillance based on random sampling. In other words, testing a proportion of a certain subgroup is less or as efficient as testing a proportion of this subgroup in each administrative unit (i.e., fever clinic or hospital).

## Notes

### Competing Interest Statement

The authors have declared no competing interest.

### Funding Statement

YL and MJ are funded by the National Institute of Health Research (UK)(16/137/109), the Bill & Melinda Gates Foundation (INV-003174), and the European Commission (101003688). PK is funded by the Royal Society (UK)(RP\EA\180004), the Bill & Melinda Gates Foundation (INV-003174), and the European Commission (101003688). SF and SC are funded through a Sir Henry Dale Fellowship jointly funded by the Wellcome Trust and the Royal Society (Grant Number 208812/Z/17/Z). MES is funded by the National Institute of Allergy and Infectious Diseases (US)(T32AI074492).
This research was partly funded by the National Institute for Health Research (NIHR) (16/137/109) using UK aid from the UK Government to support global health research. The views expressed in this publication are those of the author(s) and not necessarily those of the NIHR or the UK Department of Health and Social Care. This research is partly funded by the Bill & Melinda Gates Foundation (INV-003174). The findings and conclusions in this report are those of the author(s) and do not necessarily represent the official position of the Bill & Melinda Gates Foundation.

### Author Declarations

London School of Hygiene and Tropical Medicine

